# Re-emergent Tremor during stable posture in Parkinson’s Disease: Evidence of Pathological Beta and Prokinetic Gamma Activity

**DOI:** 10.1101/2023.08.23.23294492

**Authors:** Hao Ding, Bahman Nasseroleslami, Daniela Mirzac, Jens Volkmann, Gunter Deuschl, Sergiu Groppa, Muthuraman Muthuraman

## Abstract

Re-emergent tremor (RET) during stable posture in Parkinson’s disease (PD) is characterized as a continuation of resting tremor (RT) and is often highly therapy refractory. The pathophysiology of both RT and RET is linked to deficits in cerebello-cortical circuits and dopaminergic depletion. However, it remains unclear how these two types of tremors differ functionally. The aim of this study is to examine the differences in brain activity between RT and RET in PD, as well as to investigate the relationship between neuronal oscillatory activity, in PD tremor phenotype patients.

Forty PD patients (25 males, mean age 66.78 ± 5.03 years) and 40 age- and sex-matched healthy controls were assessed. 256 channel HD-EEG and EMG signals were recorded while the participants extended their hands against gravity. Tremor was recorded in both L-dopa ON and OFF for PD patients and mimicked by healthy controls. Coherent sources of EEG-EMG were located using beamforming technique, and information flow between different sources was estimated using time-resolved partial-directed coherence. Cross-frequency coupling (CFC) was then used to analyze the association between tremor frequency and neuronal oscillatory activity.

Under L-dopa administration, coherent sources referenced to the tremor frequency revealed differences in brain activity between RT and RET in the premotor cortex and cerebellum of PD patients, which were similar to those observed in healthy controls. However, PD patients exhibited an additional source location in the primary sensorimotor cortex. Withdrawal of L-dopa led to coherent sources being observed in the supplementary motor area and subthalamic nucleus. Furthermore, L-dopa was found to suppress the strength of connections between these coherent sources and modulate the tremor-associated beta and gamma frequency, leading to a decrease in beta power and an increase in gamma power.

The findings of this study reveal discernible neural activity patterns during resting and re-emergent tremors. In patients with PD, the primary sensorimotor cortex plays a primordial role as the exclusive source of activity involved in the generation of RET. It shows a significant amount of co-activation within the network involving the premotor areas and cerebellum, and its activation accounts for the discrepancy in tremor phenotypes. Moreover, the oscillatory neuronal responses involve pathological beta and prokinetic gamma activity alterations that are highly specific to tremor phenotypes. These peculiarities play an essential role in our understanding of tremor phenomena and its therapeutic modulation by dopamine medication or deep brain stimulation, which could specifically target the tremor- and motor-control-related pathological beta and prokinetic gamma oscillations.

## Introduction

Parkinson’s disease (PD) is characterized by a variety of symptoms, with tremor being one of the cardinal signs. Tremor is a highly heterogeneous symptom, ranging from mild to severe manifestations, and presenting as rest, postural, and kinetic tremor (Dirkx and Bologna 2022). Resting tremor (RT) in PD is commonly described as asymmetric, with a frequency of 4-6 Hz, and typically involves the hands in a pill-rolling pattern (Muthuraman et al. 2018; Lenka and Jankovic 2021). On the other hand, postural and kinetic tremor are both action tremors, manifesting during activity (Bhatia et al. 2017). The inhibition of tremor during voluntary movements is a characteristic of the RT in PD (Deuschl et al. 2012). Re-emergent tremor (RET) is a type of tremor that can occur during stable posture but also stable kinetic action in PD patients. During stable posture, it emerges after a time latency while the arms are held in a horizontal position (Lenka and Jankovic 2021). As it shares similar frequency characteristics with RT (Jankovic et al. 1999), it was proposed to be the “continuation” of resting tremor (Dirkx et al. 2018).

One of the widely accepted models of tremor pathology posits that the basal ganglia act as a “light-switch” for motor activity, initiating tremor episodes, while the cerebello-thalamo-cortical circuit functions as a “light-dimmer,” modulating tremor rhythm (Dirkx and Bologna 2022). Within this circuit, the motor cortex plays a specific role in determining tremor amplitude, whereas the thalamus and cerebellum may be important for maintaining the tremor rhythm (Helmich 2018). Previous studies have shown that there are common pathophysiological mechanisms for both RT and RET, in which the motor cortex plays an important role (Leodori et al. 2020). However, different dopamine response was found in RET (Dirkx et al. 2018). More recently, transcranial magnetic stimulation at cerebellum was found to reset RET but not RT (Helmich et al. 2021). These findings suggest that the two tremor types exhibit differences in their common pathophysiological mechanisms and involve non-dopaminergic areas such as the cerebellum during RET.

In terms of the cerebral cortex, specific regions within the cerebello-cortical network, namely the sensorimotor cortex (SMC), premotor cortex (PMC), and supplementary motor areas (SMA), have demonstrated the ability to differentiate between RT observed in Parkinson’s disease patients and mimicked tremor in healthy individuals (Muthuraman et al. 2012, 2017). Based on the frequency of tremor, cerebro-cerebral coherence analysis has revealed a widespread network connected to the M1, including the SMA and lateral PMC (Timmermann et al. 2003). While the cerebello-thalamo-cortical circuit is responsible for sustaining the tremor (Helmich 2018), periods of the tremor were found to be associated with enhanced synchronization of low beta power in the PMC (Lauro et al. 2021). These findings underscore the involvement of these motor regions in the mechanism of tremor and suggest a potential role in bridging the gap between RT and RET.

Building on prior research, this study posits that the cerebellum is implicated in cortico-subcortical communication differences between RT and RET during stable posture in PD. Thus, the study endeavours to explore brain network fingerprints and oscillatory interactions between the two tremor types in PD. To achieve this, the study analyses 256 channel – high density electroencephalography (EEG) - electromyography (EMG) data captured simultaneously from PD patients during RT and RET, as well as mimicked tremor from healthy participants. The investigation utilizes structural MRI data and EEG signal to reconstruct coherent source activity in tandem with EMG signal as reference to identify brain activity distinctions between the two tremor types. Effective connectivity analyses are also conducted to identify influential associations between these brain regions. To explore the difference of tremor oscillatory interactions between the identified source activity, this study employs cross-frequency coupling between the differentiated tremor power from cortical source regions and high-frequency power (beta, gamma) from subcortical source regions. Additionally, the study evaluates the influence of dopamine medication based on the identified region as well as its associated clinical measures. By integrating multiple datasets (EEG, EMG, MRI, and clinical measures), this study has the potential to yield significant findings on the underlying networks involved in RT and RET (Figure 1).

**Figure 1.**
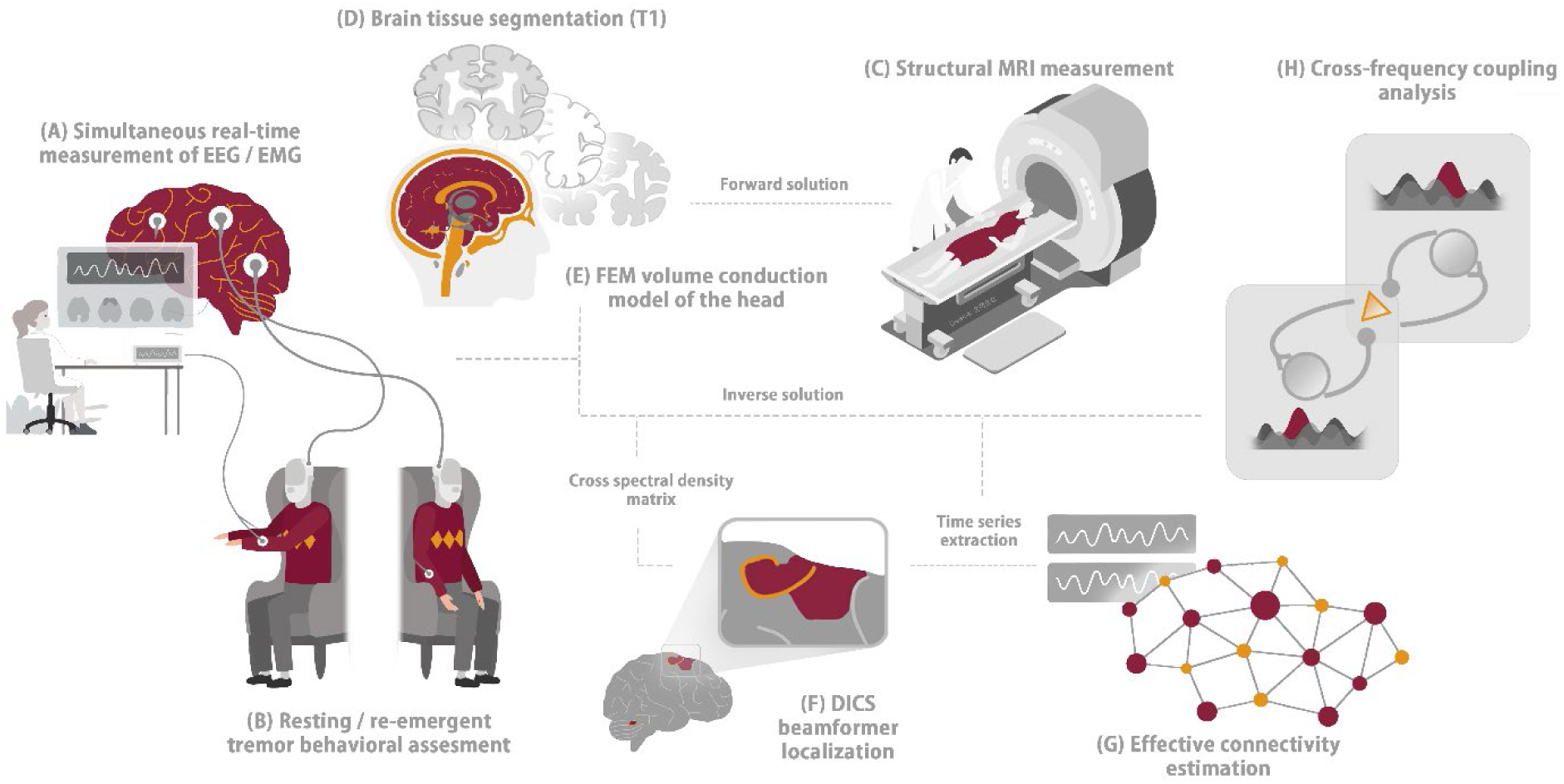
Schematic of experiment and analysis pipeline. **(A)** EEG-EMG measurements were simultaneously recorded from the participants. **(B)** PD patients performed both resting and re-emergent tremors during the measurement, while healthy participants mimicked the two tremor types. **(C)** All participants underwent structural MRI acquisition. **(D)** Brain tissue segmentation performed based on the T1 image. **(E)** Realistic head model construction performed using the FEM method. **(F)** DICS technique utilized for source localization. **(G)** Effective connectivity estimation applied to the timeseries extracted from the coherent source. **(H)** Cross-frequency coupling analysis performed on the timeseries extracted from the coherent source. PD Parkinson’s disease, EEG electroencephalography, EMG electromyography, MRI magnetic resonance imaging, FEM finite element method, DICS dynamic imaging of coherent sources.

## Results

### Participant demographics

**Table 1:**
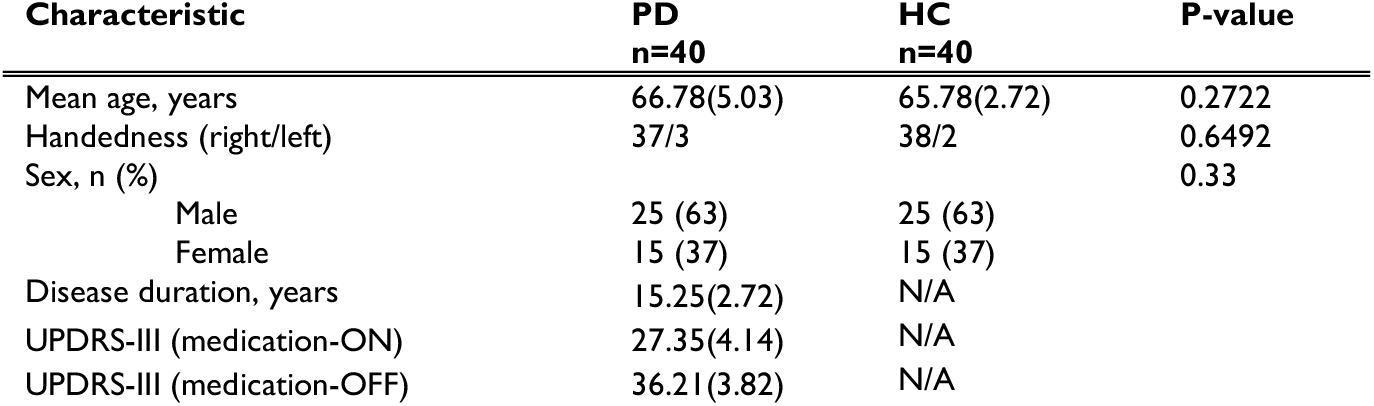
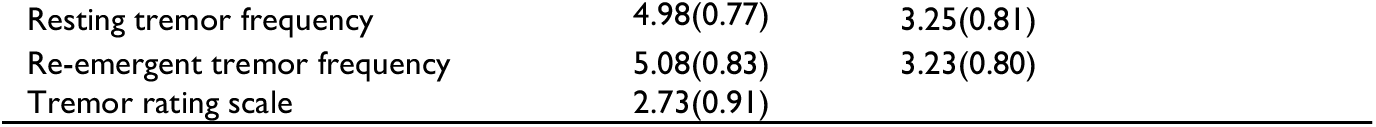
Demographics and disease characteristics. PD Parkinson’s disease, HC healthy control, (#) standard deviation.

| Characteristic | PD<br>n=40 | HC<br>n=40 | P-value |
| --- | --- | --- | --- |
| Mean age, years | 66.78(5.03) | 65.78(2.72) | 0.2722 |
| Handedness (right/left) | 37/3 | 38/2 | 0.6492 |
| Sex, n (%) |  |  | 0.33 |
| Male | 25 (63) | 25 (63) |  |
| Female | 15 (37) | 15 (37) |  |
| Disease duration, years | 15.25(2.72) | N/A |  |
| UPDRS-III (medication-ON) | 27.35(4.14) | N/A |  |
| UPDRS-III (medication-OFF) | 36.21(3.82) | N/A |  |

|  |  |  |
| --- | --- | --- |
| Resting tremor frequency | 4.98(0.77) | 3.25(0.81) |
| Re-emergent tremor frequency | 5.08(0.83) | 3.23(0.80) |
| Tremor rating scale | 2.73(0.91) |  |

In this study, a comparison was made between the demographic and clinical characteristics of the study participants belonging to the PD and control groups. The PD group exhibited a mean age of 66.78 ± 5.03 years, which was slightly higher than that of the control group (65.78 ± 2.72 years), however, this difference was not found to be statistically significant (p > 0.05). Furthermore, there was no significant difference observed in handedness between the two groups (p > 0.05). Regarding clinical characteristics, the PD group demonstrated more pronounced RT and RET frequency as compared to the mimicked RT and RET observed in the control group.

### Reconstructed source locations and the connectivity interactions under medication

Our source analysis revealed differences in brain activity between resting and re-emergent tremors in both PD patients and controls, as shown in Figure 2A. Specifically, PD patients exhibited brain activity in the primary sensorimotor cortex (PSMC), PMC, and cerebellum (CER) under L-dopa ON condition. However, upon withdrawing the medication, PD patients demonstrated source locations in two brain regions, namely the SMA and subthalamic nucleus (STN). In contrast, controls showed a similar pattern as PD patients under L-dopa ON condition, but without significant activity in the PSMC.

**Figure 2.**
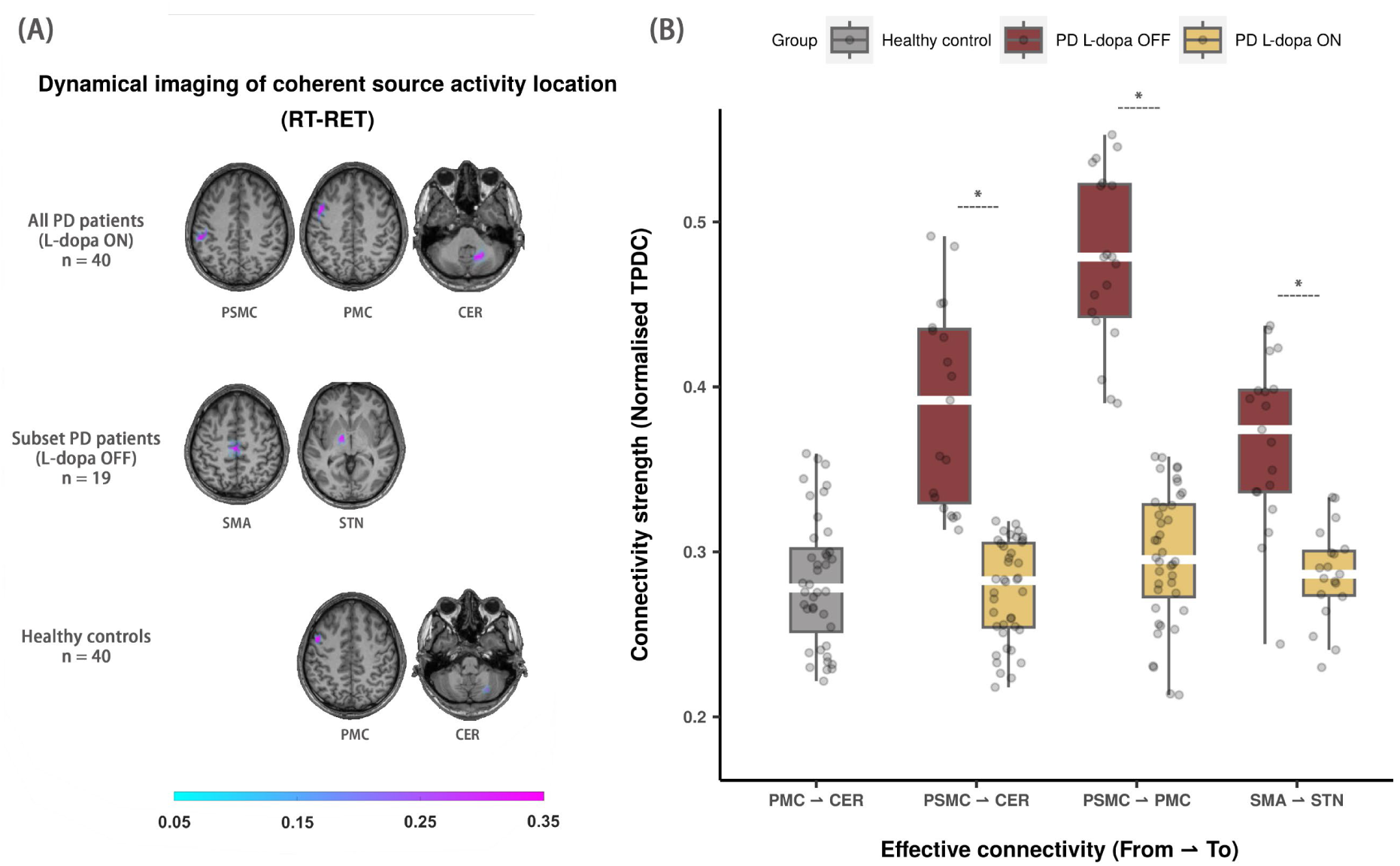
Coherent source locations and the connectivity interactions. **(A)** Reconstructed coherent source locations contrasted by resting tremor and re-emergent tremor during L-dopa ON and OFF conditions in PD patients and healthy controls. The first row shows the source locations based on all PD patients during L-dopa ON. The second row shows the source locations based on 19 PD patients who were measured during L-dopa OFF. The third row shows the source locations based on 40 healthy controls. **(B)** Boxplot representation of four unidirectional connectivity that were found to be statistically significant, along with their modifications under dopamine medication. PD Parkinson’s disease, PSMC primary sensorimotor cortex, PMC premotor cortex, CER cerebellum, SMA supplementary motor area, STN subthalamic nucleus, TPDC time-resolved partial directed coherence. RT resting tremor, RET re-emergent tremor * p < 0.05

Furthermore, connectivity analysis revealed four statistically significant unidirectional connectivity at individual tremor frequency, as depicted in Figure 2B. These connectivity pairs included PSMC ⇀ PMC, PSMC ⇀ CER, and SMA ⇀ STN among PD patients, and PMC ⇀ CER in healthy controls. Notably, PD patients exhibited a higher level of connectivity strength in L-dopa OFF compared to L-dopa ON condition, with PSMC ⇀ PMC showing the highest median compared to the other connectivity pairs (p < 0.001). After L-dopa administration, PD patients demonstrated reduced connectivity in PSMC ⇀ CER (p < 0.001), PSMC ⇀ PMC (p < 0.001), and SMA ⇀ STN (p < 0.001). However, no significant difference was observed in the connectivity pairs under L-dopa ON condition in PD patients (PSMC ⇀ CER: p > 0.05; PSMC ⇀ PMC: p > 0.05; SMA ⇀ STN: p > 0.05) and between the PMC ⇀ CER connectivity in controls (PSMC ⇀ CER: p > 0.05; PSMC ⇀ PMC: p > 0.05; SMA ⇀ STN: p > 0.05). Detailed results for comparison between L-dopa ON and OFF per significant connectivity pairs are summarized in Supplementary Table 1.

### Cross-frequency coupling and dopamine medication effects

Following the identification of significant connectivity pairs between the source locations, we conducted a cross-frequency coupling analysis to investigate the neural oscillatory interaction between cortical tremor frequency and subcortical neuronal activity, as illustrated in Figure 3. Both PD patients and healthy controls showed a tendency of a reversed relationship between beta and gamma frequency bands from subcortical regions with cortical tremor frequency. In the absence of L-dopa, PD patients exhibited significantly higher cross-frequency coupling strength at beta frequency in PSMC-CER (p < 0.001), PSMC-PMC (p < 0.001), and SMA-STN (p < 0.001), but significantly lower coupling strength at gamma frequency in PSMC-CER (p < 0.001), PSMC-PMC (p < 0.001), and SMA-STN (p < 0.001) compared to the L-dopa ON condition. In healthy controls, the cortical tremor frequency power from PMC coupled with beta frequency in CER at a lower level relative to the coupling strength at gamma frequency. No difference was observed in power-to-power CFC related to alpha frequency band. Detailed results are summarized in Supplementary Table 1 and Figure 1.

**Figure 3.**
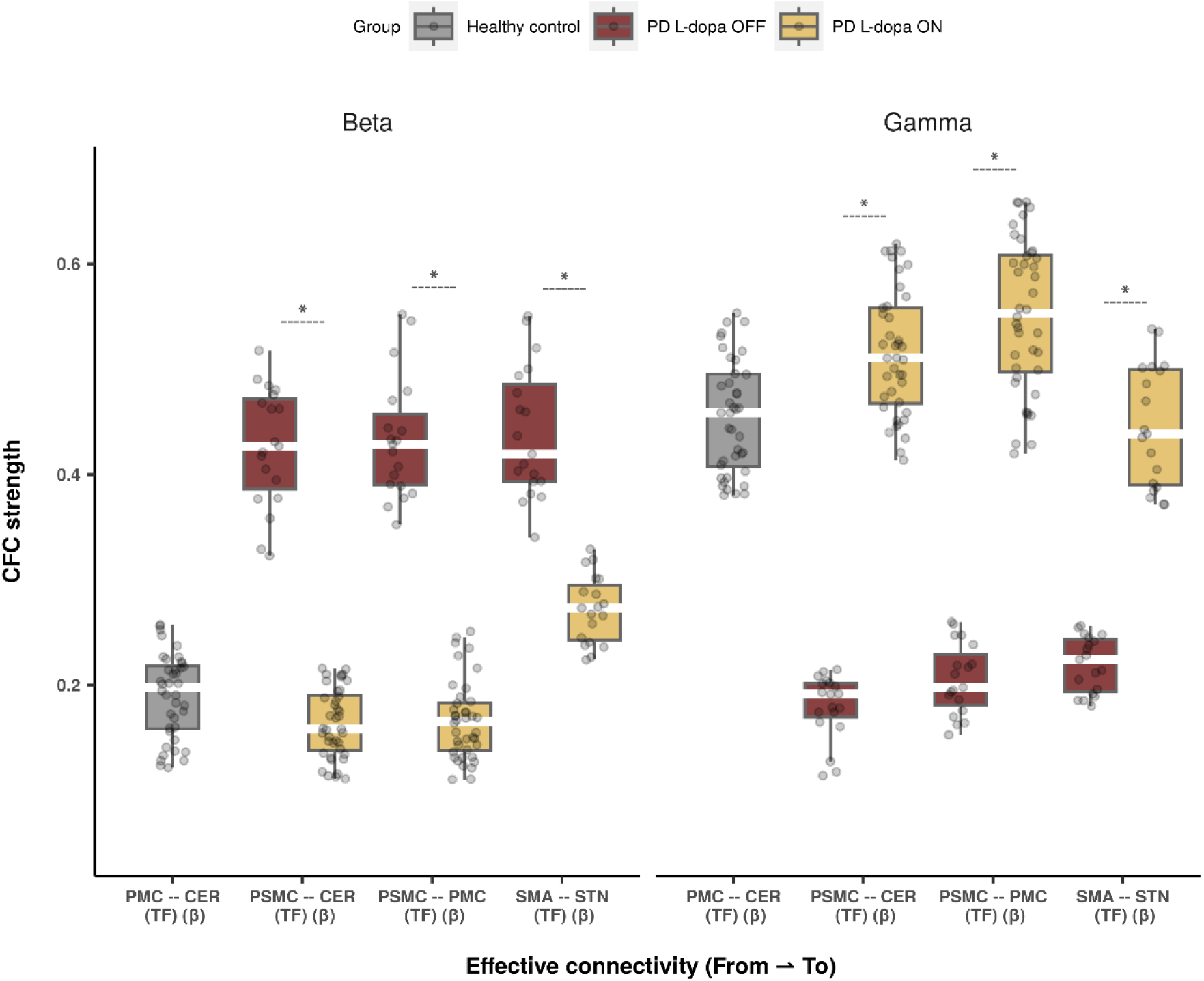
Cross-frequency coupling between tremor frequency and beta/gamma oscillations. The boxplot illustrates the strength of CFC between cortical source activity of individual tremor frequency and high-frequency bands (beta and gamma) from subcortical source under L-dopa ON and OFF conditions. PD Parkinson’s disease, RT resting tremor, RET re-emergent tremor, PSMC primary sensorimotor cortex, PMC premotor cortex, CER cerebellum, SMA supplementary motor area, STN subthalamic nucleus, CFC cross-frequency coupling, TF individual tremor frequency, β beta frequency band, γ gamma frequency band * p < 0.05

### Clinical association with the strength of cross-frequency coupling and connectivity

Finally, we investigated the associations between quantified measures and clinical evaluations including UPDRS-III and Hoehn and Yahr scale. Correlation analysis revealed a positive relationship between UPDRS-III scores and effective connectivity in PSMC ⇀ PMC (r = 0.3694, p < 0.05) during L-dopa ON condition (Figure 4A) while a negative correlation with effective connectivity in PSMC ⇀ CER (r = 0.4834, p < 0.05) during L-dopa OFF condition (Figure 4B). No statistical significance observed in other effective connectivity with UPDRS-III or Hoehn and Yahr scale. Based on these results, we further probe the correlation between neural oscillations obtained from CFC and the clinical evaluations. During L-dopa ON condition, moderate positive correlation was observed between Hoehn and Yahr scale and CFC strength at both beta (r = 0.4222, p < 0.01) and gamma (r = 0.3308, p < 0.05) frequency band in PSMC – CER (Figure 4C and 4D). Furthermore, no significant correlations were observed with clinical measures. Detailed results are summarized in Supplementary Table 2 and 3.

**Figure 4.**
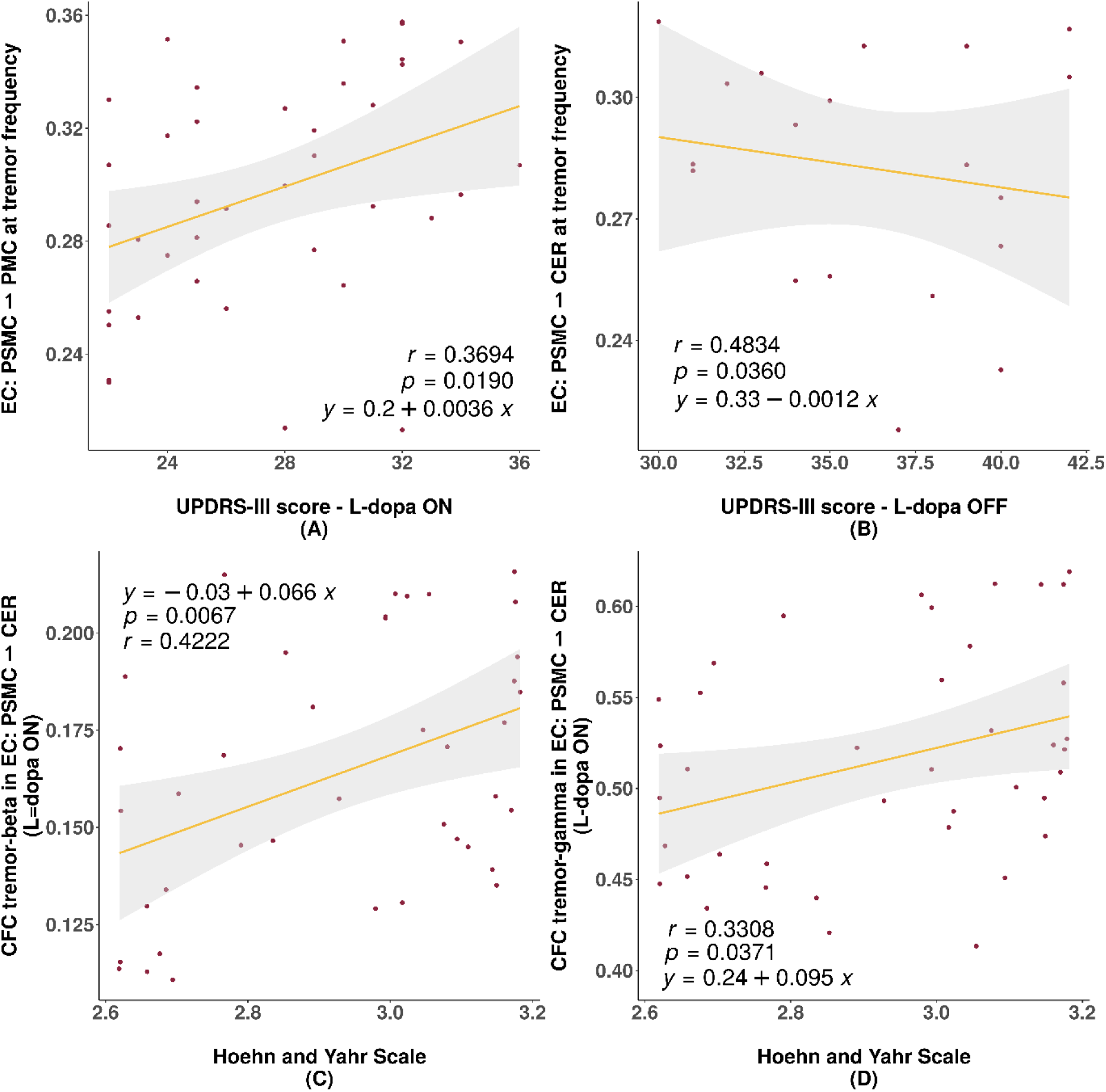
Correlation analysis between clinical measures and both the strength of cross-frequency coupling and connectivity in PD patients. **(A)** Positive correlation between effective connectivity in PSMC ⇀ PMC and UPDRS-III during L-dopa ON. **(B)** Negative correlation between effective connectivity in PSMC ⇀ CER and UPDRS-III during L-dopa OFF. **(C)** Positive correlation between Hoehn and Yahr scale and the CFC strength between cortical tremor frequency from PSMC and beta frequency from CER. **(D)** Positive correlation between Hoehn and Yahr scale and the CFC strength between cortical tremor frequency from PSMC and gamma frequency from CER. PSMC primary sensorimotor cortex, PMC premotor cortex, CER cerebellum, CFC cross-frequency coupling, UPDRS Unified Parkinson’s Disease Rating Scale, *r* Pearson correlation coefficient.

## Discussion

The present study investigated the EEG-EMG coherent source activity in resting and re-emergent tremors, focusing on the premotor and cerebellum regions in PD patients (L-dopa ON) and healthy controls. Using effective connectivity estimation, the study identified exclusive source activity in the primary sensorimotor cortex in PD patients, which may contribute to the different mechanisms of the two tremor phenotypes. Notably, without dopamine medication, PD patients exhibited a different pattern of source activity in differentiating resting and re-emergent tremors. Additionally, the study examined the power-to-power cross-frequency coupling between the source activity at tremor frequency in cortical regions and beta and gamma oscillations in subcortical regions during L-dopa ON and OFF conditions. The results revealed that dopamine medication can influence the coupling between cortical tremor frequency and subcortical beta and gamma activity. These findings imply the presence of discernable neural activity patterns distinguishing resting tremors from re-emergent tremors. It is evident that dopamine medication distinctly influences the relationship linking cortical tremor frequency with beta and gamma activities.

Previous studies have suggested that re-emergent tremor shares the same central tremor circuit as resting tremor and may be an extension or continuation of it (Dirkx et al. 2018). However, our current study identified different coherent sources between the two tremor phenotypes. Both PD patients (L-dopa ON) and healthy controls exhibited significantly difference of source activity in the premotor and cerebellum regions, which are known to play crucial roles in tremor circuits. Specifically, the premotor cortex is involved in the central representation of resting tremor, while the cerebellum plays a modulator role (Timmermann et al. 2003).

Both premotor and cerebellum were found to be involved in motor imagery (Taube et al. 2015) and have been implicated in the pathophysiology of tremor (Muthuraman et al. 2012). In the context of resting tremor, the lack of voluntary movement may cause the overactivity in the premotor cortex and cerebellum. On the other hand, re-emergent tremor may involve sensory feedback generated by repositioning the limb from a resting to an outstretched posture. This feedback could increase somatosensory input and produce reverberations within the cerebello-thalamo-cortical tremor circuit (Dirkx and Bologna 2022), potentially reducing the need for modulation in the premotor cortex and cerebellum. This could explain the lower levels of source activity observed in these regions during re-emergent tremor compared to resting tremor. Nonetheless, the present study found that connectivity from the premotor to the cerebellum in PD patients was not statistically significant, which is contradictory to the healthy controls. This might suggest a potential dysfunction of oscillatory flow between the two regions directly.

Interestingly, primary sensorimotor cortex was appeared to differentiate resting and re-emergent tremor in PD patient. This region is known responsible for sensory processing and its involvement in tremor has been reported in previous studies (Playford et al. 1992; Muthuraman et al. 2017). Effective connectivity estimation further revealed a significant association from the primary sensorimotor cortex to both the premotor cortex and cerebellum, suggesting a key role of primary sensorimotor cortex as potential modulator within the cortico-subcortical interaction in terms of the difference between resting and re-emergent tremor in PD. Furthermore, these two primary sensorimotor cortex related connections showed an association with motor symptoms during L-dopa OFF and ON conditions respectively. Thus, the primary sensorimotor cortex may play a crucial role as an exclusive modulator in the cerebello-thalamo-cortical loop in PD patients. This aligns with the reported functional reorganization of the sensorimotor cortex associated with PD (Kojovic et al. 2012), which may reflect either compensatory changes or maladaptive plasticity in response to abnormal basal ganglia activity that affects to sensorimotor and its associated cortical areas (Milardi et al. 2019). Moreover, the strength of the connectivity related to primary sensorimotor cortex decreased after the medication. One possible explanation is that L-dopa could partially normalize the connectivity of the basal ganglia motor circuit (Gao et al. 2017), leading to a reduction in the cortico-cerebellar loop. Consequently, the flow of information from the primary sensorimotor cortex to the cerebellum may have been attenuated.

In addition, the observed differences in coherent source activity during L-dopa off condition provide further support for the altered neural circuitry underlying the distinction between resting and re-emergent tremor in PD patients. Specifically, the coherent source activity exhibited a distinct pattern (SMA and STN) compared to the L-dopa ON condition (PSMC, PMC and CER) and the tremor pattern seen in healthy controls (PMC and CER). A previous study suggested that the supplementary motor area plays a crucial role in parkinsonian tremor and is coupled with other motor areas with pathologically synchronized activity (Timmermann et al. 2003). Meanwhile, the subthalamic nucleus may be involved in triggering tremor through its connectivity with the globus pallidus internus and in maintaining the tremor rhythm or both (Helmich 2018). One possible explanation for the distinct pattern during L-dopa OFF condition is that due to insufficient dopamine levels as input to the striatum, re-emergent tremor may depend less on the basal ganglia circuitry but more on the cerebello-thalamo-cortical circuit. This may also explain a previous finding by (Helmich et al. 2021) that transcranial magnetic stimulation at cerebellum can reset re-emergent tremor but not resting tremor. It might also support why re-emergent tremor is less responsive to L-dopa in some studies (Dirkx et al. 2018), as it involves non-dopaminergic areas such as the cerebellum.

Previous studies highlighted cortical regions such as primary motor cortex as a key role in controlling tremor amplitude (Leodori et al. 2020; Helmich et al. 2021), and its wide network with other cortical regions within cortico-thalamo-cerebellar loop involved in the pathophysiology of tremor in PD (Timmermann et al. 2003). In current study, cross-frequency coupling applied on the tremor power from cortical region and high frequency bands (beta and gamma) from subcortical regions revealed the neuronal interactions affected by the dopamine medication. As the time series of each source activity already represented the difference between two tremor phenotypes, our findings firstly demonstrated there is a difference of power in the frequency domain. Even though a few studies reported no difference in mean frequency between resting and re-emergent tremor (Jankovic et al. 1999; Bellows and Jankovic 2022), several studies showed a visible higher frequency and peak power of re-emergent tremor compared to resting tremor (Dirkx et al. 2018; Helmich 2018; Helmich et al. 2021). Secondly, the mimicked tremor from healthy controls exhibited CFC between tremor frequency from premotor region coupled with beta power from the cerebellum, at a relatively lower level, in contrast to gamma frequency. This suggests that there is a hypoactive gamma oscillatory modulation involved in the differential circuitry between resting and re-emergent tremor in PD. Similarly, PD patients could attain the same degree of coupling in the cortico-subcortical network through the primary sensorimotor cortex (PSMC - PMC and PSMC - CER), while under the influence of dopamine medication. This observation is consistent with previous studies on the oscillatory profile of the sensorimotor cortex in PD. It may aid in facilitating movement to counteract the anti-kinetic bias induced by the dopamine-depleted state (Rowland et al. 2015). In our dataset, a positive correlation was observed between the Hoehn and Yahr scale and the coherence of cortical-cerebellar connectivity at both beta and gamma power. This finding suggests that increased strength of coupling is associated with more severe symptoms of PD, which is consistent with the observations that coupling levels significantly increased in the absence of dopamine medication. Therefore, it is plausible that the cortico-cerebellar loop not only contributes to tremor oscillations by influencing the thalamo-cortical system (Berg and Helmich 2021) in the two tremor phenotypes, but also impacts different motor tasks and represents the overall severity of PD.

Upon withdrawal of L-dopa, there was a significant increase in the coupling strength between the SMA and STN. This distinct pattern observed in PD patients may be caused by the alteration in the basal ganglia-thalamo-cortico network, which can lead to prominent beta oscillations in the STN (Wingeier et al. 2006). In CFC, this connection (SMA-STN) with beta oscillations is strongly associated with tremor frequency, indicating the importance of beta modulation in the mechanism of resting and re-emergent tremor. Nevertheless, the coupling strength does not decline to the same extent as other regions under the L-dopa ON condition, implying that dopamine medication may has relatively limited effect on modulating tremor-related beta power between the SMA and STN regions.

One limitation of this study is that while we have found differences in the source activity, we cannot exclude the possibility that these differences may be due to variances in the anatomical distribution of the tremors. It is important to note that the cross-frequency coupling amplitude to amplitude analysis utilized in this study is unable to reveal directional information. While it has been found that tremor amplitude is associated with cortical activity as seen in previous studies, due to the constraints of the method used in this study, we were not able to reveal the directionality of the oscillatory interactions on the tremor frequency with beta and gamma. In order to address this issue, future studies could consider the utilization of other methods to examine the directional information of the oscillatory interactions. Additionally, further research could investigate the potential relationship between the anatomical distribution of the tremors and the source activity, in order to elucidate the potential role of this factor in the observed differences in source activity.

In conclusion, the current study has provided evidence for distinctive patterns of neural activity between resting and re-emergent tremors, indicating distinct mechanisms of tremor modulation within the cortico-subcortical network. During resting tremors, the primary sensorimotor cortex may act as a primordial modulator between the premotor cortex and the cerebellum, mitigating the effects of basal ganglia circuit dysfunction in the absence of voluntary movement. Conversely, in re-emergent tremors, PD patients may rely less on the basal ganglia circuit and more on the cerebello-thalamo-cortical pathway. Moreover, the oscillatory neuronal responses provide direct evidence of pathological beta and prokinetic gamma activity in patients between RT and RET. These discoveries play a fundamental role in advancing our understanding of tremor phenomena and its therapeutic modulation through the administration of dopamine medication or the application of deep brain stimulation, which could specifically target the pathological beta and prokinetic gamma oscillations related to tremor and motor control.

## Materials and methods

### Study participants and experiment procedures

This study included forty patients who were clinically diagnosed with definite Parkinson’s disease (PD) according to the London Brain Bank criteria (Hughes et al. 1992). The patients had a mean age of 66.775 ± 10.2 years and a mean disease duration of 15.25 ± 2.75 years. The general inclusion criteria required the presence of resting tremor (UPDRS items 3.17 and 3.18) with a summed score for amplitude and constancy of at least 4, postural tremor (RET) with an amplitude score of at least 2 (UPDRS item 3.15), and tremor limited to one side that appeared after at least 3 seconds while maintaining the arm outstretched. The exclusion criteria were a history of other neurological or psychiatric conditions, advanced PD (stage IV or V on the Hoehn and Yahr scale) (Hoehn and Yahr 1967), or the presence of head tremor. None of the participants had impaired cognitive function on the clinical exam, and their medications were not changed for this study. A control group of forty age- and sex-matched volunteers with normal neurological exams was also included. Prior to participation, all participants provided written informed consent, and the study procedures were approved by the institutional review board and were in accordance with the Declaration of Helsinki.

During the experiment, participants were seated in a comfortable chair in a slightly reclined position and their forearms were supported to the wrists by firm armrests. Tremors were measured using surface EMG on the forearm, while EEG was simultaneously recorded. The recording commenced with a resting state of one minute with eyes open, followed by a prompt from the experimenter to hold the hands outstretched against gravity for another minute, while keeping their eyes open and fixed on a point approximately 2 meters away. Participants diagnosed with PD, exhibiting resting tremors that persisted during posture at a similar frequency, were selected and performed 10 trials for the study.

In contrast, healthy subjects were asked to perform rhythmic extension-flexion arm movements as fast as possible to simulate hand tremors. The participants were instructed to produce the rhythmic movements in a self-paced manner, while the consistency of the movements was monitored for each subject by analyzing the EMG activity online. A frequency of at least two to five bursts per second was required, and time windows with lower frequencies were discarded. Healthy participants who could not sustain the movements for more than one minute performed the task twice.

### EEG-EMG data acquisition and preprocessing

A high-density 256-channel recording system (EGI, www.egi.com) was utilized to record EEG data. Prior to recording, the correct head net size was determined by measuring the circumference of the subject’s head. The net was then placed over the subject’s head, with the central electrode (Cz) serving as the reference electrode located at the crossing of the midline and lateral line. EMG data were simultaneously recorded using silver-silver chloride electrodes from the forearm flexors and extensors. Data acquired from the dominant hand was used for further analysis. Raw EEG and EMG signals were recorded at a sampling rate of 1000 Hz and band-pass filtered (EMG 30–200 Hz; EEG 0.05–200 Hz). Full-wave rectification of the EMG signal was performed before filtering to produce demodulated EMG (Journee 1983). Each recording was segmented into 1-second epochs, and any data segments with visible artefacts were discarded.

### Cortico-muscular coherent sources localization

To elucidate how neural electric current gives rise to the electric potentials at external sensors, a volume conduction model was estimated. This model was aimed at generating a realistic head model and contained individual electrode locations, as well as information about the geometry and conductivity of the head. The finite-element method was employed to construct the model, which comprised compartments based on individual T1 images. The resultant model facilitated accurate forward problem computations of EEG potentials in subsequent analyses.

To establish the time-frequency domain of interest, we performed time-frequency analysis using the multitaper method proposed by (Mitra and Pesaran 1999). This approach entails estimating the spectrum by multiplying the data x(*t*) with *K* distinct windows or tapers. Further details of the technique are provided elsewhere (Muthuraman et al. 2010, 2017). We used overlapping windows of 1000 ms with a time step of 50 ms, yielding an approximate frequency resolution of 1 Hz and time resolution of 50 ms. We computed an initial coherence estimate for each individual EEG electrode and then combined them to obtain a pooled coherence estimate. The individual second-order spectra were computed using a weighting scheme, and the coherence was estimated to derive the pooled estimate of the individual EEG electrodes, as suggested by (Rosenberg et al. 1989; Amjad et al. 1997). Based on the pooled time-frequency spectrum, we selected the time intervals that showed significant coherence between the EEG and EMG signals at the tremor frequency for all patients and healthy controls.

We solved the EEG inverse problem by calculating the cross-spectral density of the frequency of interest between the EEG and EMG signals, and then estimating the underlying coherent neural activity that gives rise to the electric potentials recorded on the scalp. To this end, we utilized the DICS beamforming technique (Sekihara and Scholz 1996; Gross and Ioannides 1999) which applies a spatial filter (Veen et al. 1997) to compute tomographic maps of cerebro-muscular coherence in the frequency band of interest. The analysis was conducted separately for the resting and re-emergent tremor recordings, and the difference between the two conditions was calculated. Source analysis was specifically targeted towards the cortical, subcortical, and cerebellar regions, leveraging the advantages of the high-density electrode array of EEG cap. This strategic approach guaranteed superior signal-to-noise ratios, facilitating precise identification and localization of neural sources. The spatial filter was employed across numerous voxels in these areas, assigning coherence value to each voxel. The chosen granularity for each voxel was 2 mm.

The regions of interest with the highest coherence to the EMG signal at the tremor frequency were identified as the source. Since the coherence between these regions is always 1, they were removed from the coherence matrix to identify additional coherent regions. The individual maps of the strongest cerebro-muscular coherence were normalized and averaged, then displayed on a standard MNI template brain using statistical parametric mapping toolbox. A grid was created for each subject, aligned to the Montreal neurologic institute coordinates to consider individual anatomical differences. The local maxima in the resulting maps represent the areas with the strongest coherence to the EMG signal, and these sources were extracted as time series for connectivity analysis.

### Effective connectivity estimation

By employing time-frequency connectivity analysis, we can target a specific frequency and investigate the changes in connectivity over time. To assess the directed connectivity at a particular frequency, we utilized the time-resolved partial directed coherence (TPDC) method. TPDC has been widely employed in previous studies analyzing time series data such as EEG, EMG, and MEG (Bourguignon et al. 2015; Muthuraman et al. 2020), owing to its ability to disregard indirect influences. This approach employs the dual-extended Kalman filter (Wan and Nelson 2001) to estimate the time-dependent autoregressive coefficients, which are then subjected to Fourier transformation to compute partial directed coherence (PDC) (Baccalá and Sameshima 2001). PDC between time series *x_j_* and *x_i_* at each time point can be determined by:

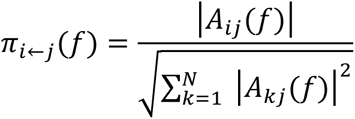

The strength of the connection in the frequency domain is evaluated using PDC based on the principle of Granger causality, where A*_ij_* represents the Fourier transformed multivariate autoregressive coefficients, and *N* is the number of pairwise connections. Following the squaring of PDC values, the resultant normalized value lies between 0 and 1. The frequency bands employed for analysis corresponded to the individual re-emergent tremor frequency. The significance level of the estimated TPDC values was calculated using a bootstrapping method (Kamiński et al. 2001), which involved the division of the original time series into non-overlapping windows, followed by random shuffling of the window order to create a new time series. This shuffled time series was fitted with a multivariate autoregressive model, and the TPDC was estimated. The shuffling process was repeated 1000 times, and the 95th percentile of the averaged TPDC value was considered as the significance threshold for all connections, with the procedure being performed individually for each patient.

### Cross-frequency coupling analysis

Cross-frequency coupling (CFC) refers to the interactions between different frequency bands, and it is believed to play a critical role in various behavioral tasks (Jirsa and Müller 2013; Jafakesh et al. 2016). Power to power is a well-known form of CFC, which demonstrates how amplitude modulations in one frequency depend on the amplitude modulations in another frequency (Davoudi et al. 2021). This coupling can be quantified using the following equation:

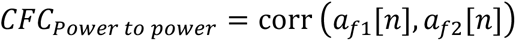

where the Pearson correlation was computed between the instantaneous amplitude of two frequency bands *f1* and *f2*. This approach has an advantage over coherence as it can identify couplings between different frequencies and within the same region. To investigate this type of coupling, the power was separately analyzed in individual RET frequency in cortical source region and alpha (8-13 Hz), beta (14-30 Hz), and gamma (31-100 Hz) frequency bands in subcortical source regions. The relationship between tremor frequency and canonical frequency bands was estimated by using a window length of 5 seconds with a 50% overlap.

### Statistical analysis

If not stated otherwise, all analyses were conducted using custom-written R scripts (version 4.2.1). The ggplot2 package was utilized to generate figures (Wickham 2016). The normality of the data was assessed using the Kolmogorov-Smirnov test. To test for differences between L-dopa conditions, paired-samples t-tests were performed. Pearson correlation was employed to determine any clinical relevance between connectivity and cross-frequency coupling outcomes. A level of statistical significance of p < 0.05 (two-tailed) was applied to test against the null-hypothesis. Due to the hypothesis-driven nature of the correlation analysis and the limited number of multiple comparisons conducted in this study, a correction method for p-values was deemed unnecessary.

## Data Availability

All data produced in the present study are available upon reasonable request to the authors

## Acknowledgements

We extend our gratitude to all participants who participated in this study. We would also like to express our gratitude to Miss Meng Shi for her invaluable advice in graphic design for the manuscript.

## Funding

This work was supported by the German Research Foundation (DFG): SFB-TRR-295 (to M.M.), MU 4354/1-1(to M.M.) and the and the Fondazione Grigioni per il Morbo di Parkinson by IUI.

## Competing interests

The authors declare that the research was conducted in the absence of any commercial or financial relationships that could be construed as a potential conflict of interest.

## Notes

### Competing Interest Statement

The authors have declared no competing interest.

### Funding Statement

This study was funded by the German Research Foundation (DFG): SFB-TRR-295 (to M.M.), MU 4354/1-1(to M.M.) and the and the Fondazione Grigioni per il Morbo di Parkinson by IUI.

### Author Declarations

Ethics commitee of medical faculty in university of Kiel gave ethical approval for this work.

## References

Amjad AM, Halliday DM, Rosenberg JR, Conway BA. An extended difference of coherence test for comparing and combining several independent coherence estimates: theory and application to the study of motor units and physiological tremor. J Neurosci Meth. 1997;73(1):69–79.

Baccalá LA, Sameshima K. Partial directed coherence: a new concept in neural structure determination. Biological cybernetics. 2001;84(6):463–74.

Bellows S, Jankovic J. Parkinsonism and tremor syndromes. J Neurol Sci. 2022;433:120018.

Berg KRE van den, Helmich RC. The Role of the Cerebellum in Tremor – Evidence from Neuroimaging. Tremor Other Hyperkinetic Movements. 2021;11(1):49.

Bhatia KP, Bain P, Bajaj N, Elble RJ, Hallett M, Louis ED, et al. Consensus Statement on the classification of tremors. from the task force on tremor of the International Parkinson and Movement Disorder Society: IPMDS Task Force on Tremor Consensus Statement. Mov Disord. 2017;33(1):75–87.

Bourguignon M, Piitulainen H, Tiège XD, Jousmäki V, Hari R. Corticokinematic coherence mainly reflects movement-induced proprioceptive feedback. Neuroimage. 2015;106:382–90.

Davoudi S, Ahmadi A, Daliri MR. Frequency–amplitude coupling: a new approach for decoding of attended features in covert visual attention task. Neural Comput & Applic. 2021;33(8):3487–502.

Deuschl G, Papengut F, Hellriegel H. The phenomenology of Parkinsonian tremor. Parkinsonism Relat D. 2012;18:S87–9.

Dirkx MF, Bologna M. The pathophysiology of Parkinson’s disease tremor. J Neurol Sci. 2022;435:120196.

Dirkx MF, Zach H, Bloem BR, Hallett M, Helmich RC. The nature of postural tremor in Parkinson disease. Neurology. 2018;90(13):e1095–103.

Gao L, Zhang J, Chan P, Wu T. Levodopa Effect on Basal Ganglia Motor Circuit in Parkinson’s Disease. Cns Neurosci Ther. 2017;23(1):76–86.

Gross J, Ioannides AA. Linear transformations of data space in MEG. Phys Medicine Biology. 1999;44(8):2081–97.

Helmich RC. The cerebral basis of Parkinsonian tremor: A network perspective. Mov Disord. 2018;33(2):219–31.

Helmich RC, Berg KREV den, Panyakaew P, Cho HJ, Osterholt T, McGurrin P, et al. Cerebello-Cortical Control of Tremor Rhythm and Amplitude in Parkinson’s Disease. Mov Disord. 2021;36(7):1727–9.

Hoehn MM, Yahr MD. Parkinsonism onset, progression, and mortality. Neurology. 1967;17(5):427–427.

Hughes AJ, Daniel SE, Kilford L, Lees AJ. Accuracy of clinical diagnosis of idiopathic Parkinson’s disease: a clinico-pathological study of 100 cases. J Neurology Neurosurg Psychiatry. 1992;55(3):181.

Jafakesh S, Jahromy FZ, Daliri MR. Decoding of object categories from brain signals using cross frequency coupling methods. Biomed Signal Proces. 2016;27:60–7.

Jankovic J, Schwartz KS, Ondo W. Re-emergent tremor of Parkinson’s disease. J Neurology Neurosurg Psychiatry. 1999;67(5):646.

Jirsa V, Müller V. Cross-frequency coupling in real and virtual brain networks. Front Comput Neurosc. 2013;7:78.

Journee HL. Demodulation of Amplitude Modulated Noise: A Mathematical Evaluation of a Demodulator for Pathological Tremor EMG’s. Ieee T Bio-med Eng. 1983;BME-30(5):304–8.

Kamiński M, Ding M, Truccolo WA, Bressler SL. Evaluating causal relations in neural systems: Granger causality, directed transfer function and statistical assessment of significance. Biol Cybern. 2001;85(2):145–57.

Kojovic M, Bologna M, Kassavetis P, Murase N, Palomar FJ, Berardelli A, et al. Functional reorganization of sensorimotor cortex in early Parkinson disease. Neurology. 2012;78(18):1441–8.

Lauro PM, Lee S, Akbar U, Asaad WF. Subthalamic-Cortical Network Reorganization during Parkinson’s Tremor. J Neurosci : Off J Soc Neurosci. 2021;41(47):9844–58.

Lenka A, Jankovic J. Tremor Syndromes: An Updated Review. Front Neurol. 2021;12:684835.

Leodori G, Belvisi D, Bartolo MID, Fabbrini A, Costanzo M, Vial F, et al. Re-emergent Tremor in Parkinson’s Disease: The Role of the Motor Cortex. Mov Disord. 2020;35(6):1002–11.

Milardi D, Quartarone A, Bramanti A, Anastasi G, Bertino S, Basile GA, et al. The Cortico-Basal Ganglia-Cerebellar Network: Past, Present and Future Perspectives. Frontiers Syst Neurosci. 2019;13:61.

Mitra PP, Pesaran B. Analysis of Dynamic Brain Imaging Data. Biophys J. 1999;76(2):691–708.

Muthuraman M, Bange M, Koirala N, Ciolac D, Pintea B, Glaser M, et al. Cross-frequency coupling between gamma oscillations and deep brain stimulation frequency in Parkinson’s disease. Brain. 2020;143(11):awaa297-.

Muthuraman M, Galka A, Deuschl G, Heute U, Raethjen J. Dynamical correlation of non-stationary signals in time domain—A comparative study. Biomed Signal Proces. 2010;5(3):205–13.

Muthuraman M, Heute U, Arning K, Anwar AR, Elble R, Deuschl G, et al. Oscillating central motor networks in pathological tremors and voluntary movements. What makes the difference? Neuroimage. 2012;60(2):1331–9.

Muthuraman M, Raethjen J, Koirala N, Anwar AR, Mideksa KG, Elble R, et al. Cerebello-cortical network fingerprints differ between essential, Parkinson’s and mimicked tremors. Brain J Neurology. 2017;141(6):1770–81.

Muthuraman M, Schnitzler A, Groppa S. [Pathophysiology of tremor]. Nervenarzt. 2018;89(4):408–15.

Playford ED, Jenkins IH, Passingham RE, Nutt J, Frackowiak RSJ, Brooks DJ. Impaired mesial frontal and putamen activation in Parkinson’s disease: A positron emission tomography study. Ann Neurol. 1992;32(2):151–61.

Rosenberg JR, Amjad AM, Breeze P, Brillinger DR, Halliday DM. The Fourier approach to the identification of functional coupling between neuronal spike trains. Prog Biophysics Mol Biology. 1989;53(1):1–31.

Rowland NC, Hemptinne CD, Swann NC, Qasim S, Miocinovic S, Ostrem JL, et al. Task-related activity in sensorimotor cortex in Parkinson’s disease and essential tremor: changes in beta and gamma bands. Front Hum Neurosci. 2015;9:512.

Sekihara K, Scholz B. Generalized Wiener estimation of three-dimensional current distribution from biomagnetic measurements. Ieee Transactions Bio-medical Eng. 1996;43(3):281–91.

Taube W, Mouthon M, Leukel C, Hoogewoud HM, Annoni JM, Keller M. Brain activity during observation and motor imagery of different balance tasks: An fMRI study. Cortex. 2015;64:102–14.

Timmermann L, Gross J, Dirks M, Volkmann J, Freund H, Schnitzler A. The cerebral oscillatory network of parkinsonian resting tremor. Brain. 2003;126(1):199–212.

Veen BDV, Drongelen WV, Yuchtman M, Suzuki A. Localization of brain electrical activity via linearly constrained minimum variance spatial filtering. Ieee T Bio-med Eng. 1997;44(9):867–80.

Wan EA, Nelson AT. Dual extended Kalman filter methods. Kalman filtering and neural networks. 2001;123.

Wickham H. ggplot2: Elegant Graphics for Data Analysis. 2016; Available from: https://ggplot2.tidyverse.org

Wingeier B, Tcheng T, Koop MM, Hill BC, Heit G, Bronte-Stewart HM. Intra-operative STN DBS attenuates the prominent beta rhythm in the STN in Parkinson’s disease. Exp Neurol. 2006;197(1):244–51.

