## Supplemental Figure and Tables for "Re-emergent Tremor during stable posture in Parkinson’s Disease: Evidence of Pathological Beta and Prokinetic Gamma Activity"

### 1 Title

### 2 Authors and affiliations

Hao Ding[1,2], Bahman Nasseroleslami[2], Daniela Mirzac[3], Jens Volkmann[1], Gunter Deuschl[4], Sergiu Groppa[3], Muthuraman Muthuraman[1]

1 Department of Neurology, University hospital Würzburg, Würzburg, Germany
2 Academic Unit of Neurology, Trinity College Dublin, the University of Dublin, Dublin, Ireland
3 Department of Neurology, University Medical Center of the Johannes Gutenberg-UniversityMainz, Mainz, Germany
4 Department of Neurology, UKSH, Christian-Albrechts-University Kiel, Kiel, Germany

### Supplementary Material

| **Quantified measures contrasts between L-dopa ON and OFF** | **p value** |
|---|---|
| **Effective connectivity** | |
| PSMC⇁CER | **<0.001** |
| PSMC⇁PMC | **<0.001** |
| SMA⇁STN | **<0.001** |
| **Power-to-Power CFC at alpha** | |
| PSMC-CER | 1 |
| PSMC-PMC | 1 |
| SMA-STN | 1 |
| **Power-to-Power CFC at beta** | |
| PSMC-CER | **<0.001** |
| PSMC-PMC | **<0.001** |
| SMA-STN | **<0.001** |
| **Power-to-Power CFC at gamma** | |
| PSMC-CER | **<0.001** |
| PSMC-PMC | **<0.001** |
| SMA-STN | **<0.001** |

Table 1: Statistical analysis between L-dopa ON and OFF condition effective connectivity and power-to-power CFC.

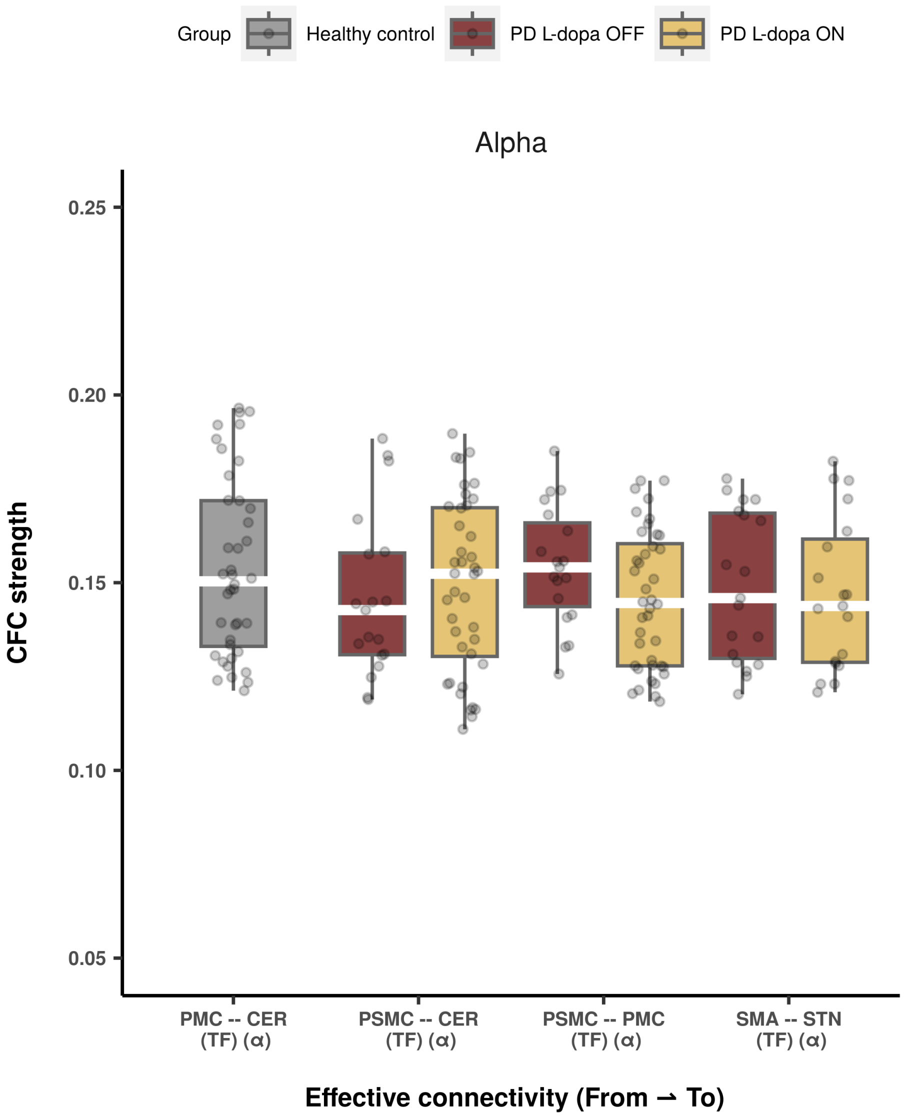

Figure 1: Cross-frequency coupling between tremor frequency and alpha oscillations. The boxplot illustrates the strength of CFC between cortical source activity of individual tremor frequency and high-frequency bands (alpha) from subcortical source under L-dopa ON and OFF conditions. PD Parkinson's disease, RT resting tremor, RET re-emergent tremor, PSMC primary sensorimotor cortex, PMC premotor cortex, CER cerebellum, SMA supplementary motor area, STN subthalamic nucleus, CFC cross-frequency coupling, TF individual tremor frequency, $\alpha$ alpha frequency band

| **Group** | **Effective connectivity** | **Hoehn and Yahr scale** | **UPDRS-III (L-dopa ON)** | **UPDRS-III (L-dopa OFF)** |
|---|---|---|---|---|
| | | **Correlation coefficient $r$/p-value** | | |
| Parkinson's disease patients | PSMC$\rightharpoonup$PMC (L-dopa ON) | -0.1115/0.4933 | **0.3694/0.019** | 0.0737/0.7642 |
| | PSMC$\rightharpoonup$CER (L-dopa ON) | 0.0433/0.7907 | 0.0748/0.6464 | -0.1606/0.5113 |
| | SMA$\rightharpoonup$STN (L-dopa ON) | 0.2461/0.3098 | -0.2473/0.3075 | 0.1394/0.5692 |
| | PSMC$\rightharpoonup$PMC (L-dopa OFF) | 0.0636/0.796 | -0.1643/0.5015 | -0.3108/0.1953 |
| | PSMC$\rightharpoonup$CER (L-dopa OFF) | 0.2983/0.2148 | 0.1858/0.4463 | **0.4834/0.036** |
| | SMA$\rightharpoonup$STN (L-dopa OFF) | 0.2886/0.2308 | -0.2625/0.2776 | -0.0236/0.9237 |
| Healthy controls | PMC$\rightharpoonup$CER | -0.1979/0.2209 | 0.07/0.668 | 0.0476/0.8466 |

Table 2: Correlation summary between TPDC and clinical measures.

| Power-to-power CFC | Hoehn and Yahr scale | UPDRS-III (L-dopa ON) | UPDRS-III (L-dopa OFF) |
|---|---|---|---|
| | Correlation coefficients / p-value | | |
| **CFC at beta (L-dopa ON)** | | | |
| Tremor frequency-beta PSMC-PMC | -0.0091/0.9556 | 0.1147/0.4811 | -0.2654/0.2721 |
| Tremor frequency-beta PSMC-CER | **0.4222/0.0067** | -0.2009/0.2139 | -0.0815/0.7403 |
| **CFC at beta (L-dopa OFF)** | | | |
| Tremor frequency-beta PSMC-PMC | 0.072/0.7695 | 0.0336/0.8913 | 0.1959/0.4214 |
| Tremor frequency-beta PSMC-CER | 0.1352/0.581 | 0.0868/0.7237 | -0.174/0.4761 |
| **CFC at gamma (L-dopa ON)** | | | |
| Tremor frequency-gamma PSMC-PMC | -0.1332/0.4127 | 0.0516/0.7519 | 0.2052/0.3993 |
| Tremor frequency-gamma PSMC-CER | **0.3308/0.0371** | -0.1479/0.3623 | -0.0537/0.8272 |
| **CFC at gamma (L-dopa OFF)** | | | |
| Tremor frequency-gamma PSMC-PMC | -0.0799/0.745 | -0.0075/0.9758 | 0.0266/0.914 |
| Tremor frequency-gamma PSMC-CER | -0.1053/0.668 | 0.1167/0.6343 | -0.018/0.9416 |
| **CFC at alpha (L-dopa ON)** | | | |
| Tremor frequency-alpha PSMC-PMC | 0.1229/0.4501 | -0.1246/0.4436 | 0.0479/0.8457 |
| Tremor frequency-alpha PSMC-CER | -0.0634/0.6975 | -0.0477/0.7702 | 0.0125/0.9596 |
| **CFC at alpha (L-dopa OFF)** | | | |
| Tremor frequency-alpha PSMC-PMC | -0.1068/0.6496 | 0.0022/0.7817 | 0.3694/0.0676 |
| Tremor frequency-alpha PSMC-CER | -0.1115/0.9095 | 0.0681/0.6144 | -0.4279/0.8968 |

Table 3: Correlation summary between power-to-power cross frequency coupling at neural oscillation and clinical measures during L-dopa ON and OFF conditions.
